# Migraine Disability, Pain Catastrophizing, And Headache Severity Are Associated With Evoked Pain And Targeted By Mind-Body Therapy

**DOI:** 10.1101/2021.07.05.21259849

**Authors:** Samuel R. Krimmel, Michael L. Keaser, Darrah Speis, Jennifer A. Haythornthwaite, David A. Seminowicz

## Abstract

Meta-analysis suggests migraine patients are no more sensitive to experimentally evoked pain than healthy controls. At the same time, studies have linked some migraine symptoms to Quantitative Sensory Testing (QST) profiles. Unfortunately, previous studies associating migraine symptoms and QST have important methodological shortcomings, stemming from inappropriate statistics, small sample sizes, and frequent use of univariate statistics for multivariate research questions. In the current study we seek to address these limitations by using a large sample of episodic migraine patients (n=103) and a multivariate analysis that associates pain ratings from many thermal intensities simultaneously with 12 clinical measures ranging from headache frequency to sleep abnormalities. We identified a single dimension of association between QST and migraine symptoms that relates to pain ratings for all stimulus intensities and a subset of migraine symptoms relating to disability (Headache Impact Trauma 6 and Brief Pain Inventory interference), catastrophizing (Pain Catastrophizing Scale), and pain severity (average headache pain, Brief Pain Inventory severity, and Short Form McGill Pain Questionnaire 2). Headache frequency, allodynia, affect, and sleep disturbances were unrelated to this dimension. Consistent with previous research, we did not observe any difference in QST ratings between migraine patients and healthy controls. Additionally, we found that the linear combination of symptoms that related to QST were modified by mind-body therapy. These results suggest that QST has a selective relationship with pain symptoms even in the absence of between-subjects differences between chronic pain patients and healthy controls.

**Highlights:**

- Experimentally evoked pain ratings have been linked to migraine symptoms, though there are methodological shortcomings
- We find evoked pain ratings are related to disability, pain catastrophizing, and pain severity, but not to headache frequency, affect, nor sleep disturbances
- Evoked pain and symptoms relate even in the absence of pain sensitivity differences between patients and healthy controls
- Mind-body therapy altered symptoms that related to evoked pain
- Experimentally evoked pain should be used to study specific ensembles of symptoms, even when pain ratings do not differ between patients and healthy controls

## Introduction

Experimentally evoked pain ratings determined through Quantitative Sensory Testing (QST) have been used to investigate migraine pathophysiology for several decades. Many studies have examined if QST is pathological in migraine, with some work showing elevated pain ratings relative to healthy controls [8,32,33], and other studies indicating no difference [4,31,39]. Meta-analysis finds a lack of evidence for abnormalities in evoked pain sensitivity for migraine, at least for certain pain modalities [27]. These mixed/null findings from between-subjects studies on QST and migraine could be used to argue that experimentally evoked pain sensitivity is not a good avenue to investigate migraine pathophysiology. However, the absence of pain sensitivity differences between migraineurs and healthy controls does not address whether experimentally evoked pain measures are related to symptoms in migraine, as this is a within-subjects question.

In fact, within-subjects analyses have shown an association between QST measures and migraine patients’ affective and headache symptoms [3,28]. Such studies often examine the relationship between a single clinical variable and several QST measures. However, disorders like migraine display many symptoms beyond frequent painful headaches, including sleep disturbances [19], elevated pain catastrophizing [25], and affective abnormalities [6]. It is unclear whether experimentally evoked pain relates to all migraine symptoms, or rather to specific ensembles. This is important to understand, because experimentally evoked pain cannot be used to probe symptoms it is not associated with. While in theory it should be possible to synthesize the existing migraine symptom/QST literature, there are important limitations with previous studies including poor statistical practices (e.g., no multiple comparisons correction), low statistical power, and improper univariate analyses that treat chronic pain symptoms as independent of all other symptoms, which is untrue [1,2,13,20].

Multivariate analysis is a better approach for understanding how QST and migraine symptoms relate. One such approach is Canonical Correlation Analysis (CCA). Instead of performing correlations between two variables, CCA identifies associations between two domains of variables [17]. CCA has gained renewed interest in the neuroimaging field [41] and has been used to successfully link demographic characteristics and pathology with biological variables [21,26,35,43]. In the current study we associate clinical symptoms and pain ratings simultaneously in over 100 episodic migraine patients recruited for a clinical trial assessing enhanced mindfulness-based stress reduction (MBSR+) treatment of headaches [34]. We identified a single dimension of covariance between evoked pain and clinical symptoms, showing that evoked pain and clinical symptoms are related, at least in episodic migraine. This dimension was associated with all pain ratings and a subset of clinical features, primarily those related to disability, pain severity, and pain related cognition. We also found that this association was modified by MBSR+, indicating a potential mechanism of this non-pharmacologic therapy. Moreover, we did not observe any difference in evoked pain ratings between migraine subjects and healthy controls, indicating that QST measures can indeed be associated with chronic pain symptoms even in the absence of a between-subjects difference with healthy controls.

## Methods

Episodic migraine patients were recruited as part of a previously described clinical trial assessing MBSR+ treatment of migraine [34]. Briefly, participants were 18-65 years old and met the ICHD-3 criteria for episodic migraine with or without aura for more than a year [16]. Participants with a history of mindfulness training, severe or unstable psychiatric symptoms, and/or opioid use were excluded. All patients provided written informed consent in accordance with University of Maryland Baltimore and Johns Hopkins University standards. The study protocol was approved by the Johns Hopkins School of Medicine and the University of Maryland Baltimore Institutional Review Boards. After excluding subjects with any missing data, 103 subjects were available for analysis before any therapy had occurred (92 female, average age = 37.4, s.d. = 12.3). A subset of patients were randomized to receive either stress management for headache (SMH) or enhanced mindfulness based stress reduction (MBSR+). 97 subjects had requisite data to allow for assessment of MBSR+ effects on clinical canonical variate scores (47 SMH, 50 MBSR), and 93 for MBSR+ effects on QST canonical variate scores (43 SMH, 47 MBSR+). 37 healthy control subjects were also collected and matched based on age, sex, education, body mass index, and race to the first cohort of recruited migraine subjects (32 female, average age = 38.5, s.d. = 13.3).

### Clinical Assessment

A total of 12 clinical variables were used for analyses. We used the total scores for questionnaires where applicable. Disability was captured using the Brief Pain Inventory (BPI) interference score [10] and with the Headache Impact Test 6 (HIT-6; [29]). Headache diaries were used to determine the number of headache days (NHA, see [34]). Pain severity was captured by the average self-reported intensity of headache from the headache diaries [34], the Brief Pain Inventory pain severity scores [10], and the Short Form McGill Pain Questionnaire 2 (SFMPQ2; [11]). Allodynia was assessed using Allodynia Symptom Checklist scores (ASC; [18]). Sleep quality was captured with the Pittsburg Sleep Quality Index (PSQI; [7]). Pain related cognition was assessed using the Pain Catastrophizing Scale (PCS; [38]). Somatization and somatic symptom severity were captured using the Patient Health Questionnaire 15 (PHQ-15; [23]). Affective symptoms were measured using the GAD-7 to assess anxiety [37] and the Patient Health Questionnaire 9 (PHQ-9) to measure depression [22].

### Quantitative Sensory Testing (QST)

We used a 30 x 30 mm ATS probe (Medoc Pathway model, ATS, Medoc Advanced Medical Systems Ltd., Ramat Yishai, Israel). We presented a set of 19 pseudorandom thermal stimulations using a simple ramp up and hold design, and had participants rate intensity and unpleasantness using a 0-10 scale (0: no pain; 10: maximum pain imaginable). Only intensity was used for this analysis. For each of these 19 stimulations, the baseline temperature was 32°C and the temperature rose at a rate of 1.6°C a second until reaching the target temperature followed by a 6 second duration stimulus with 6 seconds between stimulations. The stimulus order was 40, 42, 44, 47, 41, 49, 41, 45, 48, 39, 49, 45, 48, 47, 45, 44, 43, 49, 47°C. All stimulation was done on the left volar surface of the participants’ forearms.

### Univariate Association of Symptoms and QST

We examined the association between QST ratings through Pearson correlation analysis between all 19 pain ratings using a false discovery rate (FDR) multiple comparisons correction [5]. We repeated this same procedure for clinical symptoms by correlating all 12 variables with one another, again with FDR correction.

### Canonical Correlation Analysis

In the current analysis, we sought to test whether there is a relationship between migraine symptoms and evoked pain ratings. Correlating all clinical measures with all QST measures would require correction for more than 200 comparisons. This approach would also treat all symptoms as independent of one another, and all pain ratings as independent of other pain ratings. To address these limitations, we used Canonical Correlation Analysis [17]. CCA is a method for identifying multivariate relationships between two domains of data measured on the same set of individuals. With CCA, a pair of canonical variates is created (one per data domain) through a linear combination of input variables so that the correlation between canonical variates is maximized (canonical correlation, R_c_). CCA assumes limited multicollinearity, so we performed a Principal Components Analysis (PCA) on clinical and QST data separately to create orthogonal components before performing CCA. We used all components in the model, so PCA was therefore not used as a dimension reduction technique. In CCA, the maximum number of canonical correlations is given by the smaller the number of variables in each domain, which in our analysis was 12. Each pair of canonical variates is orthogonal to the preceding canonical variates and has a smaller canonical correlation.

To mitigate overfitting, we used a L2 norm regularized CCA [15]. To optimize the model, we selected lambda values for clinical and QST data based on reducing overfitting. We determined optimal regularization parameters by computing the first canonical correlation in held out data from 10 fold cross validation with 10 replacements (i.e., 100 models were tested in total). Based on held out data, we selected a lambda value of 2 for the questionnaires and 3 for the QST. This resulted in average R_c_ values of .18 for testing data and .24 for training data, indicating some overfitting, but much less than without regularization (difference greater than .5). We used a permutation test to assess significance of the CCA by constructing 10,000 bootstraps with replacement [12]. For each bootstrap, we calculated R_c_ values for every canonical correlation and compared this null distribution against the true R_c_ values using a one-sided t-test.

Because we first performed PCA, the interpretation of the rotation matrix is challenging, and we therefore transformed the CCA weights back into their original space using the CCA model weights and the rotation matrix. We were therefore able to use the CCA model weights in the original clinical and QST spaces. Applying these weights to clinical and QST data yielded clinical and QST canonical variates respectively. We did not use these reconstructed weights directly for interpretation and instead used these weights to extract canonical variates over time. To interpret CCA results we used canonical loadings, which were created by performing Pearson correlation between canonical variates and their respective model inputs (e.g., QST values), using an FDR approach to correct for multiple comparisons.

### Effect of Treatment on Canonical Variate Scores

The majority of subjects were enrolled in a clinical trial assessing MBSR+ treatment of headaches and were randomized to receive either MBSR+ or SMH. This clinical trial found that MBSR+ reduced headache frequency and disability more than SMH [34]. Building off these previous findings, we sought to determine if there were treatment effects on the identified canonical variate scores. We tested this using a linear mixed effects model with fixed effects for treatment condition (MBSR+ or SMH), session (baseline, 10 weeks, or 20 weeks) and their interaction; we treated subjects as a random effect; the dependent variable was canonical variate score for QST or clinical symptoms.

## Results

### Effect of Migraine on QST Pain Ratings

We tested for effects of disease status on QST pain ratings using a linear mixed effects model with fixed effects for stimulus intensity, disease status (migraine or healthy), and an interaction term, along with a random effect for subject. We did not observe an effect of disease status (β=0.5, [95% CI -1.8 to 2.8], p=0.6), nor an interaction between disease status and stimulus intensity on pain ratings (β=-0.007, [95% CI -0.06 to 0.04], p=0.8, n=37 healthy and 103 migraine). These results suggest that migraine patients were no more sensitive to pain than healthy controls (Fig 1).

**Fig 1.**
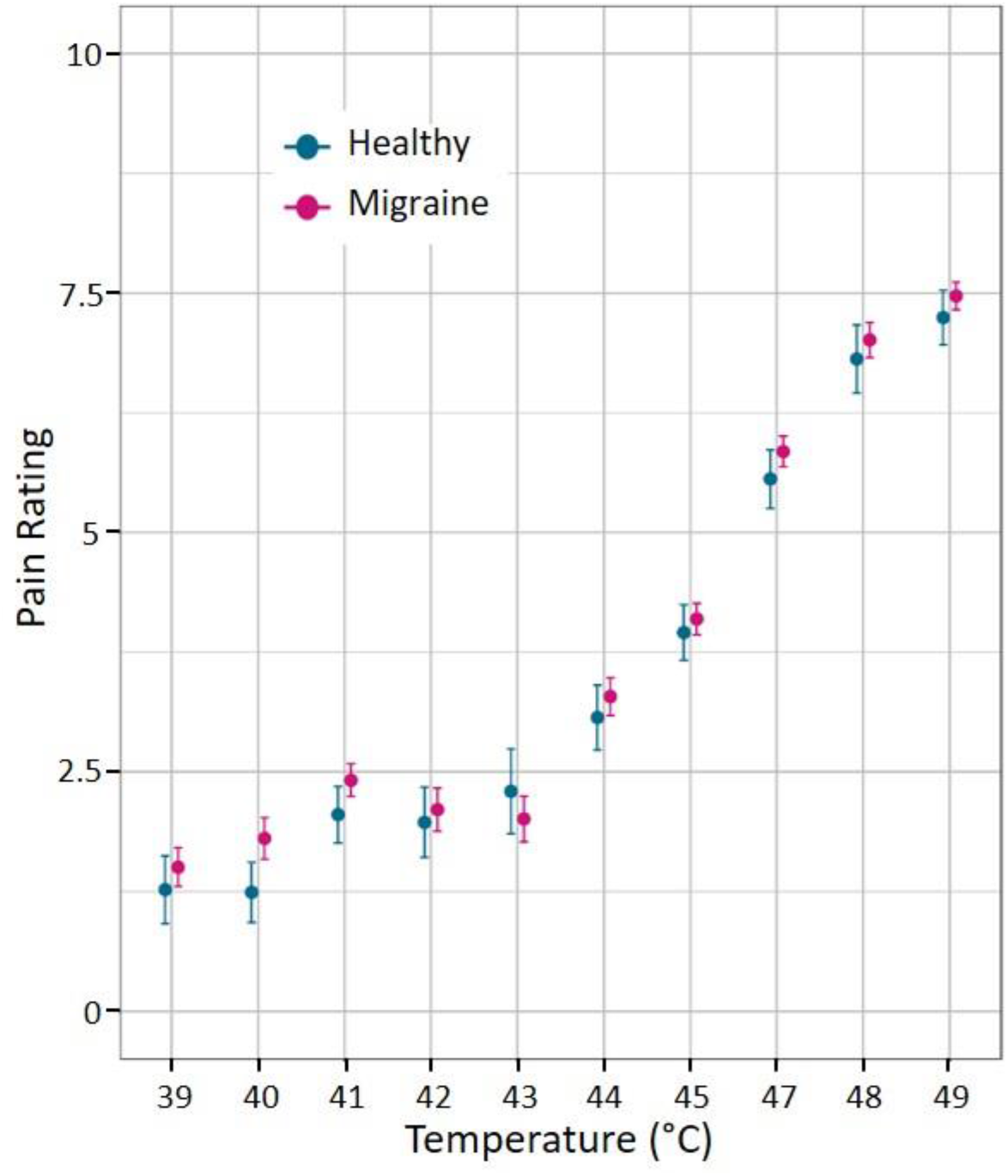
Migraine patients were not hypersensitive to noxious thermal stimuli applied to the forearm. Plot shows mean and standard error of the mean pain intensity rating (0-10) at each temperature for migraine patients (n=103) and healthy controls (n=37).

### Both Clinical Symptoms and QST are Interrelated

Using a univariate approach, we tested if pain ratings from QST were related to one another and found that every temperature rating correlated with pain ratings at all other temperatures (Fig 2A), indicating that QST data are completely interrelated. We then tested if clinical symptoms are related to one another and found that symptoms correlated with one another, but in a much sparser manner than pain ratings (Fig 2B). SFMPQ-2 scores and BPI-interference were most likely to be associated with other clinical symptoms whereas PSQI and PCS were the least likely. These analyses indicate that neither clinical symptoms nor QST responses should not be treated as independent entities for analysis.

**Fig 2.**
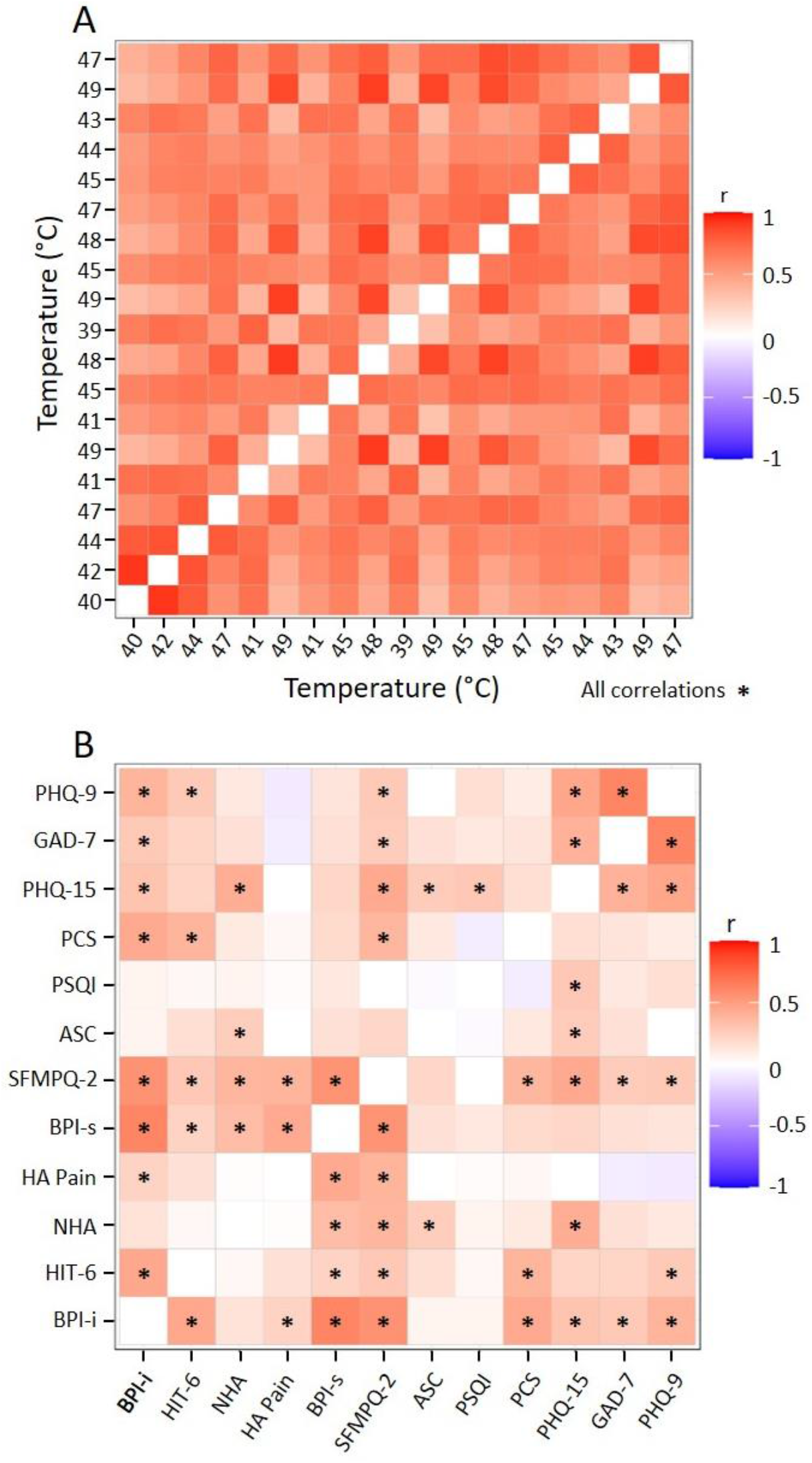
Neither migraine symptoms nor pain ratings are independent of one another. A) Correlation matrix showing relationship between pain ratings across stimulation temperatures in migraine patients (n=103). All correlations were significant. Temperature is listed in experimental order. B) Correlation matrix showing relationship between symptoms (n=103). r=Pearson correlation coefficient; see Methods: Clinical assay for symptom abbreviations; *=p<0.05, FDR-corrected for multiple comparisons.

### CCA Identifies a Dimension of Association Between QST and Clinical Symptoms

To examine the relationship between QST and clinical symptoms we used a regularized CCA on principal component scores of QST and clinical data. A total of 12 canonical correlations were created, and we found that the first canonical correlation was significant (R_c_=0.24, N=103, p<0.05, Fig 3 A,B). We only used this first canonical correlation in further CCA analysis. This canonical correlation was comprised of two scores per participant: one from the canonical variate for QST and the other from the canonical variate for clinical data.

**Fig 3.**
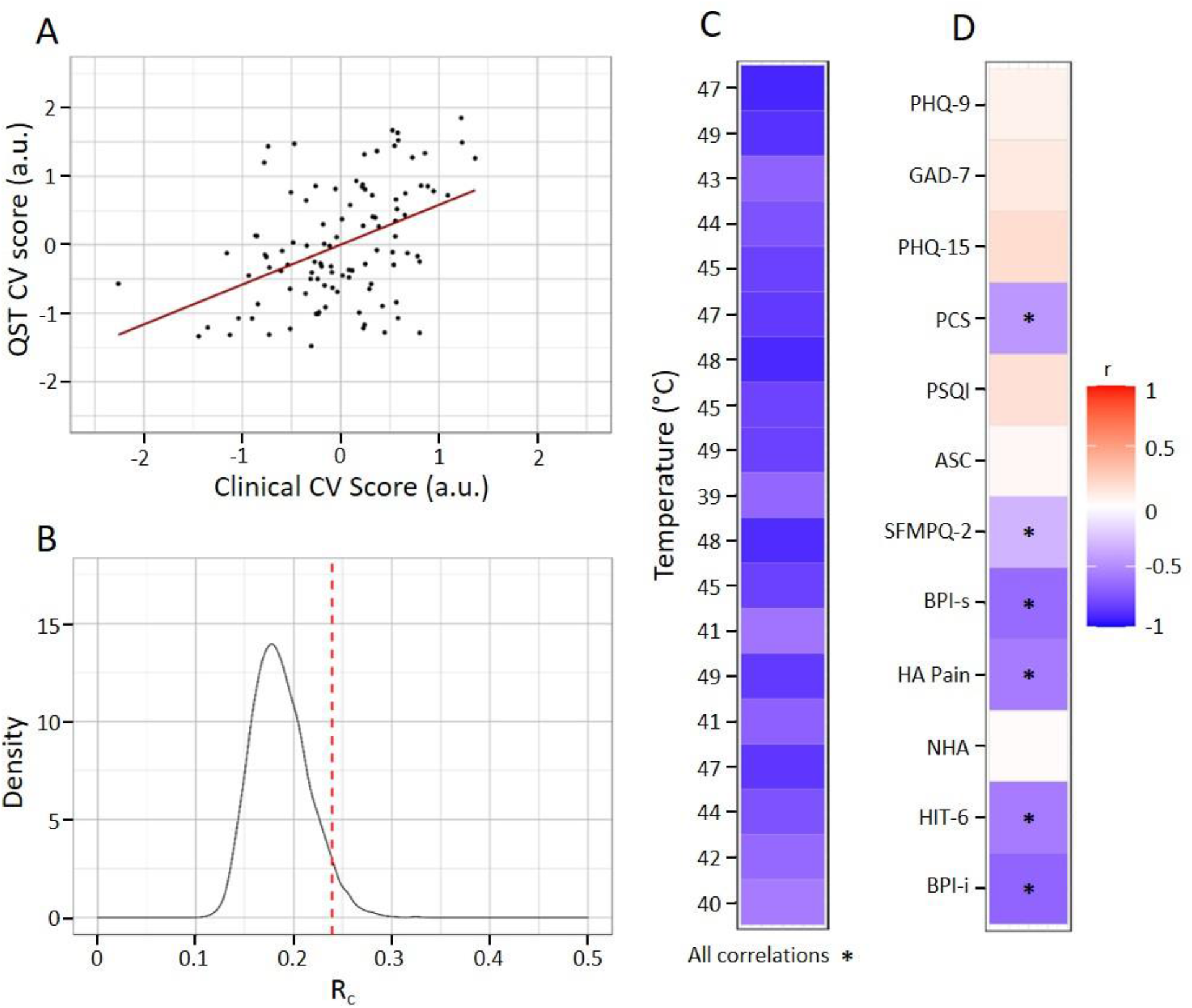
Using multivariate analysis to associate clinical features and pain ratings to thermal stimulation reveals a pain sensitivity dimension of covariance. A) First canonical correlation (R_c_) value. B) Permutation test for significance for first R_c_ value (shown in red) relative to 10,000 permutations. C) Correlation between pain ratings and QST canonical variate. All correlations were significant and in the same direction. Temperature is listed in experimental order. D) Correlation between clinical symptoms and clinical canonical variate. The significant dimension of association related to all pain ratings and to a subset of clinical features relating to pain severity, disability, and catastrophizing. Hence we refer to it as a pain sensitivity dimension. QST= quantitative sensory testing; CV=canonical variate; r=Pearson correlation coefficient; see Methods: Clinical assay for symptom abbreviations; *=p<0.05, FDR-corrected for multiple comparisons.

### Interpretation

To facilitate interpretation, we correlated canonical variate scores with the original input data, creating canonical loadings. After multiple comparisons correction, we found that all pain ratings were correlated with the QST canonical variate score in the same direction (Fig 3C). For clinical symptoms, we found that brief pain inventory severity, brief pain inventory interference, HIT-6, mean headache pain, PCS, and SFMPQ-2 were correlated with clinical canonical variate scores in the same direction (Fig 3D, Table 1). Therefore this dimension was most associated with measures related to pain severity, disability and pain-related cognition, and was not associated with affective symptoms, the frequency of headaches, sleep disturbances, somatic disturbances, nor allodynia. Importantly, the correlation pattern for non-significant clinical parameters was notably different from significant clinical parameters, where all non-significant clinical parameters had a small positive correlation (or almost zero correlation value) with the clinical canonical variate. This indicates that the null effects were not from simply missing a significance cutoff.

**Table 1.**
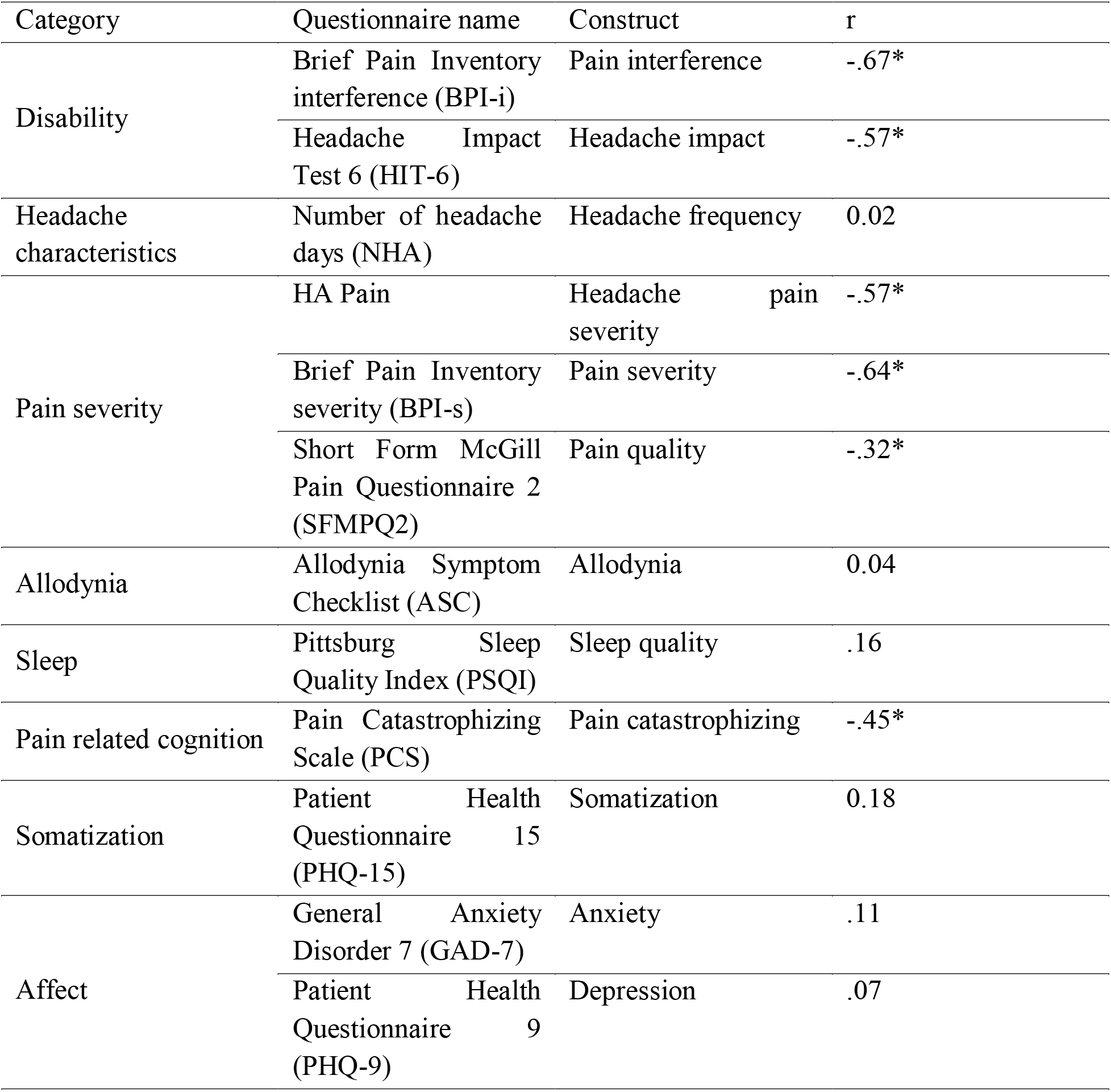
Canonical loadings for clinical canonical variate. Clinical canonical variate scores were correlated with clinical data to create canonical loadings. Clinical features were organized into categories to facilitate interpretation. Disability, pain severity, and pain related cognition were the clinical categories with significant canonical loadings. Headache characteristic, allodynia, sleep, somatization, and affect were all unrelated to clinical canonical variate scores. r=Pearson correlation coefficient; *=p<0.05, FDR-corrected for multiple comparisons.

We interpret this first dimension as a pain sensitivity dimension given that all pain ratings were correlated to canonical variate scores. Importantly, all canonical variates were standardized to be mean zero. Patients who were more pain sensitive (i.e. higher pain ratings) had negative QST canonical variate values (i.e. below average scores). Patients with negative clinical canonical variate scores had more severe values for the six clinical parameters that were weighted in the model (e.g., higher pain catastrophizing, more disability, and greater clinical pain).

### Sensitivity Dimension Change with Treatment

We tested for effects of time and treatment on canonical variate scores for symptoms and QST using a linear mixed effects model with fixed effects for treatment (MBSR+ or SMH), session (baseline, 10 weeks, 20 weeks) and an interaction effect, along with a random effect for subject. To extract these canonical variate scores, we used the CCA model weights described above, but instead used raw behavioral and clinical data (as opposed to standardized data). By not using standardization, all values were negative, and more negative values (i.e. further from zero) were for more pain sensitive subjects than were less negative values (i.e. closer to zero). We tested two separate linear mixed effects models, one for clinical canonical variate and one for QST. For clinical canonical variate scores, we found a significant treatment by time effect for clinical canonical variate scores for 10 weeks (β=0.94, [95% CI 0.04 to 1.85], p<0.05) and 20 weeks (β=1.2, [95% CI 0.3 to 2.1], p<0.05) with MBSR+ patients having less negative canonical variate scores over time relative to the active control condition (Fig 4). There was no effect of treatment on clinical canonical variate scores (β=-0.18, [95% CI -1.11 to 0.7], p=0.70). There was also no effect of time at 10 weeks (β=0.23, [95% CI -0.42 to 0.87], p=0.49), nor at 20 weeks (β=0.61, [95% CI -0.04 to 1.26], p=0.07) on clinical canonical variate scores. We did not observe any interaction between treatment and session for QST canonical variate scores at 10 weeks (β= 0.5, [95% CI -0.3 to 1.3], p=.2) nor at 20 weeks (β=0.65, [95% CI -0.1 to 1.4], p=.1). There was no effect of treatment on QST canonical variate scores (β=-0.28, [95% CI -1.16 to 0.59], p=0.5). There was also no effect of time at 10 weeks (β=0.005, [95% CI -0.55 to 0.56], p=0.9), nor at 20 weeks (β=0.08, [95% CI -0.48 to 0.63], p=0.8) on QST canonical variate scores.

**Fig 4.**
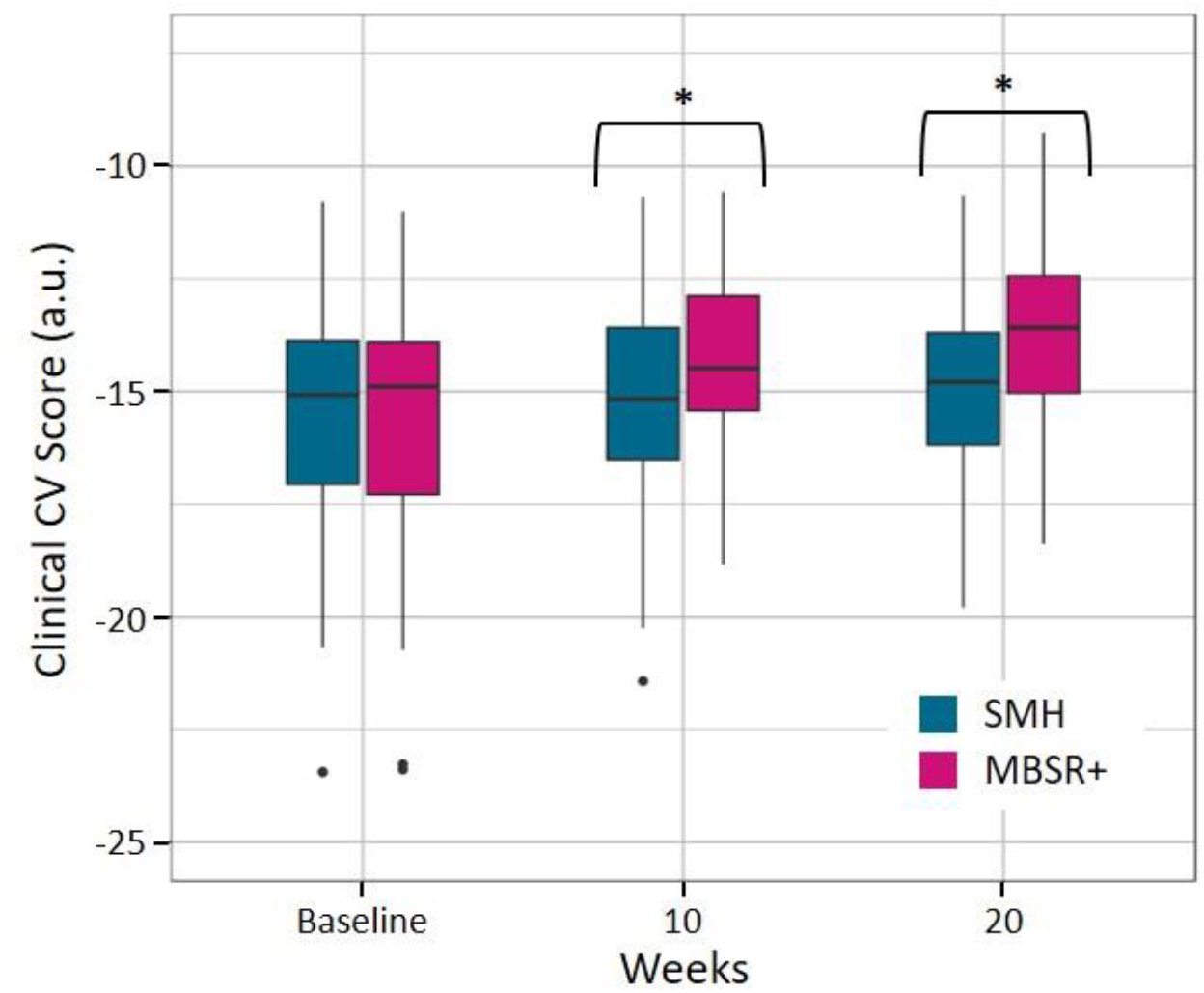
Clinical canonical variate scores are modified by therapy. Compared to patients randomized to SMH (n=47), patients randomized to MBSR+ (n=50) had clinical canonical variate scores move closer to zero over therapy. Unstandardized clinical canonical variate scores were all negative, with more negative values having more severe symptoms. Therefore, MBSR+ moved away from the more symptom-severe pole. SMH=stress management for headache; MBSR+=enhanced mindfulness-based stress reduction. *=p<0.05 for the treatment by time effect.

## Discussion

In the current study we used CCA to identify a dimension of association between experimentally evoked pain ratings and chronic pain symptoms in over 100 episodic migraineurs. This dimension related to all QST pain ratings and a subset of clinical features including pain severity, disability, and pain related cognition. Previous studies linking QST and chronic pain symptoms have been limited through analytic shortcomings such as univariate analyses, small sample sizes, and lack of multiple comparisons corrections; our approach addresses these limitations by using a large multivariate analysis that allows symptoms to be interrelated. Additionally, we found that an ensemble of clinical symptoms related to QST were modified by mind-body therapy, indicating a potential mechanism for this innovative treatment.

We interpreted the significant dimension of association as a pain sensitivity dimension because QST canonical variate scores correlated with all pain ratings in a similar manner. However, clinical canonical variate scores did not associate with all clinical variables and instead were only associated with indices of the severity of pain (mean headache pain, BPI severity, and SFMPQ-2) and response to pain such as disability (HIT-6 and BPI interference) and pain related cognition (PCS). It was not associated with affective symptoms, allodynia, nor with sleep disturbances. And surprisingly, clinical canonical variate scores did not associate with headache frequency, which is the most common target of clinical trials for migraine. This finding suggests that experimentally evoked pain is most likely to be associated with disability and clinical pain severity in migraine. Therefore, only certain illness features can be examined using standard evoked pain studies, and such techniques may not provide insight into other important symptoms. Our findings also inform ongoing debates about the utility of QST in chronic pain [30,40] and show that QST can provide insight into aspects of the chronic pain experience. Importantly, the strength of the canonical correlation was fairly small (final model R_c_ was 0.24), which suggests that complementary approaches are needed to better understand migraine symptom variability. Consistent with this, our group recently identified a much larger canonical correlation with migraine symptoms when using resting-state functional MRI connectivity in this same sample [21].

For a variety of reasons, many chronic pain studies rely on univariate analysis to associate pain symptoms and QST. For instance, researchers often associate clinical variables with various QST metrics, or use a linear model framework to investigate how clinical features interact with noxious stimulation to influence pain. These approaches suffer from a foundational limitation, because they treat clinical variables as independent of one another, an assumption that is not merited [1,2,12,19,23 and Fig 2B]. Our multivariate approach shows an alternative strategy where each symptom is not treated as a unique and separable element of the chronic pain experience. This allowed for the identification of an ensemble of symptoms that together related to experimentally evoked pain ratings.

The literature on evoked pain differences for migraine is mixed, with some studies showing that migraineurs are more sensitive, less sensitive, or no different than healthy controls [4,25,31– 33,39]. Consistent with a meta-analysis of QST in migraine [27], we did not observe patient hyper-sensitivity from thermal pain applied to the forearm. However, our analysis does show a QST/symptom relationship even in the absence of a between-subjects difference between patients and controls. Therefore, between-subjects differences (i.e. chronic pain patients vs. healthy controls) in QST should not be a prerequisite to analyze the within-subjects relationship between QST and pain and other clinical symptoms. In other words, even in studies where patients and controls have equivalent pain ratings, those QST data should still be used to understand variability of certain clinical features.

We found that the non-pharmacologic therapy MBSR+ influenced clinical canonical variate scores, which supplements the finding from the original clinical trial demonstrating that MBSR+ effectively reduces the frequency of headaches and headache related disability [34]. Specifically, clinical canonical variate scores become less severe with therapy (i.e. less negative), whereas the effect of treatment was not significant for QST canonical variate scores. These findings show that an ensemble of symptoms relating to experimentally evoked pain is targeted by mind-body therapy. Due to the ensemble’s relationship with evoked pain, it could serve as a potential endpoint for clinical trials assessing treatments that modify evoked pain, as is the case with MBSR [42].

The present study is not without its limitations. First, there is always the danger of over-fitting in CCA, which will create a model that fails to generalize. In the current study, we observed a small difference between the final model and the held out testing data from cross validation (average R_c_ difference of 0.06). Therefore, it is unlikely that over-fitting is a major cause for concern in the current analysis. A second limitation was the use of as single pain modality. In this study we only used heat pain for QST and did not use multiple sensory modalities. The results may therefore only apply to experimentally evoked heat pain, and not to other experimentally evoked pain responses. However, previous work indicates a very large association between mechanical, cold, and heat modalities [36], so it is likely that our results would generalize to additional pain modalities.

In conclusion, we reveal a dimension of association between experimentally evoked pain and clinical symptoms in migraine. This dimension was related to pain sensitivity and was associated primarily with clinical measures of disability, pain severity, and pain catastrophizing, but not affective symptoms nor the frequency of headaches. These findings indicate that the common use of evoked pain in laboratory settings is a valid approach for studying migraine symptoms and encourages similar investigations into other chronic pain symptoms. Additionally, we show that MBSR+ targets symptoms that relate to evoked pain. What remains unclear from our work is the neural mediators in the QST/symptom relationship. Future work should use mediation analysis, and possibly multivariate mediation analysis [9,14], to understand neural properties that underlie this relationship.

## Data Availability

Data is available upon request.

## Acknowledgments

NCCIH/NIH R01 AT007176 to DAS. The authors declare no conflicts of interest.

